# Cerebrospinal fluid findings in patients with idiopathic Normal pressure hydrocephalus – findings from a cohort study

**DOI:** 10.1101/2025.11.14.25340207

**Authors:** Sagar Poudel, Sakoon Saggu, Alfonso Fasano, Deepa Dash, Aparna Wagle Shukla, Ajay Garg, Ashish Dutt Upadhyay, Naveet Wig, Roopa Rajan, Animesh Das, MR Divya, Manjari Tripathi, Achal K. Srivastava, SS Kale, P Sarat Chandra, Ashish Suri, Pramod K Pal, Hrishikesh Kumar, Deepti Vibha, Rajesh Kumar Singh, Jasmine Parihar, Ranveer Singh Jadon, Ved Prakash Meena, Bindu Prakash, Arunmozhimaran Elavarasi

**Affiliations:** Department of Medicine, All India Institute of Medical Sciences, New Delhi, India; Department of Neurology, All India Institute of Medical Sciences, New Delhi, India; Department of Biomedical Sciences, Humanitas University, Milan, Italy; IRCCS Humanitas Research Hospital, Milan, Italy; Department of Clinical Neurological Sciences, University of Western Ontario, London, Canada; Department of Neurology, Fixel Institute for Neurological Diseases, University of Florida, Gainesville, Florida, United States of America; Department of Neuroimaging and Interventional Neuroradiology, All India Institute of Medical Sciences, New Delhi, India; Department of Biostatistics, All India Institute of Medical Sciences, New Delhi, India; Department of Neurosurgery, All India Institute of Medical Sciences, New Delhi, India; Department of Neurology, National Institute of Mental Health and Neurosciences, Bengaluru, India; Movement Disorders Program, Institute of Neurosciences, Kolkata, India

## Abstract

**Introduction:** Normal Pressure Hydrocephalus (NPH) is a treatable cause of gait disturbance, cognitive impairment, and urinary incontinence in older adults. The cerebrospinal fluid tap test (CSF-TT) is used as an ancillary test in the evaluation of idiopathic NPH (iNPH). However, there are no systematic studies on the correlation between CSF opening pressures and biochemical parameters with response to the CSF-TT and shunt responsiveness. CSF proteins are known to be mildly elevated in a few patients with iNPH. Periventricular hyperintensities are known to be correlated with intracranial pressure.

**Methods:** We compared CSF opening pressure and biochemical parameters between the CSF-TT responders (atleast 1 point reduction in modified Rankin scale 24 hours after CSF-TT) and non-responders. We also compared these parameters between those with and without MRI periventricular hyperintensities. MRI characteristics—including disproportionately enlarged subarachnoid-space hydrocephalus, periventricular white matter changes, Evans index, callosal angle, and cerebral infarcts—were also compared between patients with elevated versus normal CSF protein levels.

**Results:** CSF-TT responders had significantly higher CSF opening pressures (p = 0.04) compared to non-responders, while there were no differences in CSF biochemical parameters. Among MRI features, the callosal angle was significantly lower in patients with higher CSF protein levels (p = 0.02).

**Conclusion:** Higher CSF opening pressure within the diagnostic range was associated with CSF-TT responsiveness, while routine CSF biochemistry was not predictive. Elevated CSF protein was associated with a narrower callosal angle, highlighting the value of integrating pressure and imaging features in iNPH evaluation.

## Introduction

Normal pressure hydrocephalus is a disorder of characterized by a triad of gait disturbance, cognitive dysfunction, and urinary disturbance that responds to a Ventriculoperitoneal shunt or Lumbo-peritoneal shunt. The CSF tap test (CSF-TT) has been considered a crucial ancillary investigation prior to shunt surgery.^1,2^ Additionally, CSF findings, such as opening pressure, cellularity, and routine CSF biochemistry, are routinely performed in patients being evaluated with a large-volume CSF tap. Although the phrase ‘Normal pressure hydrocephalus’ implies that the pressure is normal, what constitutes normal is not precisely defined. It is known that the pressure may be higher in responders.^3^ Though elevated opening pressures may be suggestive of secondary causes of hydrocephalus, the range of normal pressures in idiopathic NPH(iNPH) is not clearly defined. The Japanese guidelines consider a diagnosis of probable INPH at an opening pressure cutoff of <200 mm of H2O.^4^ On the other hand, the Relkin guidelines^5^ consider a cutoff of <245 mm H2O. Several experts^3^ are of the opinion that the CSF pressure in INPH is not really ‘normal’ and have even called for a change in terminology to ‘chronic hydrocephalus’ instead of INPH. It is known that the CSF pressure waves often show numerous B waves, thereby signaling a tendency to intermittently high CSF pressures, and these patients have a better outcome than those without B waves.^6,7^ It is possible that there is a CSF pressure continuum, with individuals having pressures above a particular range exhibiting a better response to the CSF-TT. Periventricular white matter hyperintensities, commonly seen in INPH, likely reflect transependymal CSF flow and ischemic changes, and prior studies have shown that specific imaging features correlate strongly with CSF-TT results and may serve as predictors of shunt effectiveness.^8^ Some studies report higher than normal protein levels in up to 38% patients with iNPH.^9^ However, the studies on CSF proteins in iNPH are conflicting.^10,11^ CSF protein levels and biomarker analysis are recognized as potential predictors of shunt responsiveness and may provide valuable prognostic information in INPH.^12^

In this exploratory study, we compared CSF opening pressures and biochemical profiles between CSF-TT responders and non-responders, as well as between shunt responders and non-responders. We also report the comparison of CSF findings in those with and without periventricular white matter changes on imaging and compare the imaging findings between those with normal and elevated CSF proteins.

## Methods

In this ambispective cohort study of patients, we recruited consecutive patients who had hydrocephalus on MR imaging and features suggestive of iNPH, such as a reduced callosal angle and DESH with dilated sylvian fissures. These patients underwent a detailed history and clinical examination. The diagnosis of probable iNPH was made clinico-radiologically. These patients underwent the CSF-TT. The methodology has been described in detail elsewhere^13,14^ Patients underwent assessment at baseline and 24 hours after a 30-50 mL CSF-TT as per the institutional protocol.^13^ They were offered VP shunting, based on clinico-radiologic criteria, irrespective of the findings of the CSF-TT or CSF parameters. Patients who had at least one point improvement on the modified Rankin scale 24 hours following the CSF-TT were classified as tap responders. Those with a similar improvement at 24 weeks after VP shunt insertion were classified as shunt responders. The study was designed and reported as per the STROBE reporting guidelines for observational studies.

## Statistical analysis

Data were collected and managed using REDCap (Research Electronic Data Capture), a secure, web-based application designed to support data capture for research studies hosted by the All India Institute of Medical Sciences (AIIMS), New Delhi, India. Data were exported and cleaned using Microsoft Excel (Microsoft Corp., Redmond, WA, USA). Descriptive statistics, including frequencies and percentages, were used to describe categorical variables. Mean and standard deviations were used for continuous variables with a normal distribution, while medians and interquartile ranges were used for non-parametric data. For comparisons between groups, an independent samples t-test was used for normally distributed data, and the Mann–Whitney U test was used for non-normally distributed data. Dichotomous variables were compared using the Chi-squared test or Fisher’s exact test. ROC curve analysis was used to evaluate the ability of the CSF opening pressure to predict shunt responsiveness. The area under the curve (AUC) and the 95% confidence interval were computed, and the optimal cutoff was determined using the Youden index. Sensitivity and specificity were calculated at the chosen threshold. All statistical analyses were performed using Stata version 17 (StataCorp, College Station, TX, USA). A p-value of <0.05 was considered statistically significant.

## Results

We screened 48 patients with clinical or imaging features of iNPH and recruited a total of 40 patients who had probable iNPH and underwent CSF-TT during the period from 2019 to 2024.^13,14^ We report the comparison of the various CSF parameters between CSF-TT responders and non-responders in Table 1. The mean opening pressure was significantly higher in responders compared to non-responders. Other CSF biochemical parameters, including cell count, protein, glucose, and the CSF-to-blood glucose ratio, did not differ significantly between groups.

**Table 1:** Comparison of CSF findings between CSF tap test responders and non-responders.

| Parameter | CSF TT responder (N=18) | CSF TT non-responder (N=22) | P value |
| --- | --- | --- | --- |
| Opening pressure mean(SD) (n=27) | 17.7 ( 4.3) | 13.4 ( 5.7) | 0.04 |
| Cell count/mm<sup>3</sup> median (IQR) (n=40) | 0 (0-5) | 0 (0-5) | 0.35 |
| CSF Protein mg/dL median (IQR) (n=39) | 52 (40-55) | 45.5 (25-64) | 0.40 |
| CSF Glucose mg/dL, mean (SD) (n=39) | 71.7 (20.1) | 72.8 (16.2) | 0.86 |
| Ratio of CSF: Blood glucose mean(SD) (n=32) | 0.6 (0.1) | 0.6 (0.1) | 0.37 |

The comparison of these parameters between shunt responders (n=15) and non-responders (n=9) is reported in Supplementary Table 1. There was a trend towards higher CSF opening pressures in the shunt responders [18(15-21) vs 10(6-20) cm H2O] as compared to shunt non responders; however, this difference was not statistically significant. CSF protein levels were significantly higher in shunt responders compared with non-responders.

ROC analysis (Figure 1) of CSF opening pressure as a predictor of tap responsiveness yielded an area under the ROC curve (AUROC) of 0.74 (95% CI: 0.54–0.94), indicating moderate discriminatory ability. The optimal cutoff, identified by the Youden index, was ≥18 cm H_2-_O, yielding a sensitivity of 66.7% and specificity of 83.3%. We also found that one patient had an opening pressure of 28 cm H2O. However, the rest of the clinico-radiologic profile fitted into a diagnosis of normal-pressure hydrocephalus. The ROC analysis (supplementary figure 1) of CSF opening pressure as a predictor of shunt responsiveness had an AUROC of 0.71 (0.4-1).

**Figure 1.**
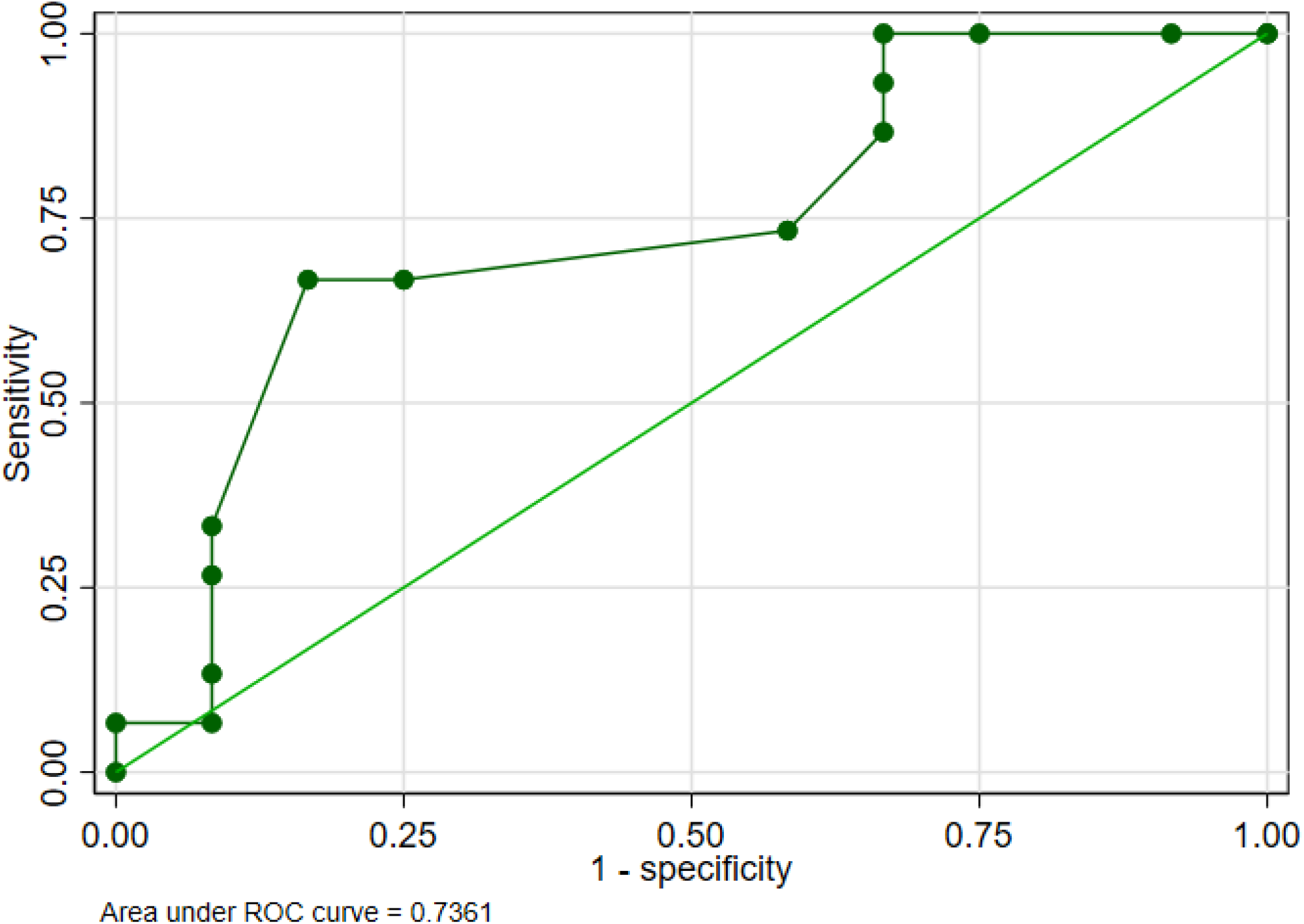
Receiver operating characteristic (ROC) curve for CSF opening pressure in predicting CSF-TT responsiveness

We compared the CSF findings between patients with and without MRI periventricular hyperintensities. (Table 2). There were no significant differences in the CSF pressures or biochemical parameters between those with and without periventricular hyperintensities on MRI.

**Table 2:** Comparison of patients with and without periventricular T2/FLAIR hyperintensities on MRI Brain.

|  | periventricular hyperintensity present n=34 | periventricular hyperintensity absent n=5 | P value |
| --- | --- | --- | --- |
| Opening pressure mean(SD) (n=27) | 15.9 (5.2) | 15.3 (6.9) | 0.83 |
| Cell count/mm<sup>3</sup> median (IQR) (n=40) | 0 (0-5) | 10 (0-35) | 0.13 |
| CSF Protein mg/dL median (IQR) (n=39) | 51.5 (34-64) | 40 (40-52) | 0.77 |
| CSF Glucose mg/dL, mean (SD) (n=39) | 73.5 (18.1) | 64.2 (14.0) | 0.28 |
| Ratio of CSF: Blood glucose mean(SD) (n=32) | 0.6 (0.1) | 0.5 (0.2) | 0.17 |

We also investigated the relationship between CSF protein levels and MRI features (Table 3). There were no differences in the proportion of those with hydrocephalus, periventricular white matter changes, DESH, widened temporal horns, dilation of the third ventricle, or Evans ratio between those with normal CSF proteins and elevated protein levels. Silent infarcts were more commonly found in the lower CSF protein group; however, the difference was not statistically significant. Patients with higher CSF protein had a significantly reduced callosal angle compared to those with normal protein levels (p = 0.02).

**Table 3:** Comparison of patients with normal and raised CSF proteins.

|  | CSF protein ≤45 mg/dL n=17 | CSF protein >45 mg/dL n=22 | P value |
| --- | --- | --- | --- |
| Hydrocephalus n(%) | 17 (100%) | 22 (100%) | - |
| Periventricular white matter changes n(%) | 20(90.9%) | 14 (82.4%) | 0.64 |
| DESH n(%) | 19 (86.4%) | 16(94.1%) | 0.62 |
| Widened temporal horns n(%) | 21 (95.5%) | 17 (100%) | >0.99 |
| Dilatation of the third ventricle n(%) | 21 (95.5%) | 17 (100%) | >0.99 |
| Bowing of corpus callosum n(%) | 16 (72.7%) | 13 (76.5%) | >0.99 |
| Infarcts n(%) | 6 (27.3%) | 1 (5.9%) | 0.11 |
| Evans ratio mean (SD) | 0.36 (0.03) | 0.36 (0.02) | 0.75 |
| Callosal angle mean (SD) | 80.2±18.0 | 67.2±16.5 | 0.02 |

## Discussion

This study provides valuable insights into the correlation between CSF opening pressure and response to the CSF-TT. Our findings suggest that opening pressure may have a predictive value for CSF-TT responsiveness, whereas routine CSF biochemistry did not differ significantly between these groups. Prior reports have highlighted that CSF pressure in iNPH is not strictly normal, with fluctuations such as B waves indicating intermittent intracranial hypertension, which is associated with better shunt outcomes.^6,7^ Our results add support to the concept that a higher baseline pressure, while still within diagnostic thresholds (<200 mm H_2_O in Japanese guidelines, <245 mm H_2_O in Relkin criteria), may identify patients more likely to benefit from shunting.^4,5^ We also had one patient with an opening pressure slightly above these thresholds. We now know from physiologic testing that CSF pressure is not uniform throughout and has peaks and pressure waves during the course of the day. Idiopathic NPH may be a continuum, with some patients experiencing intraday fluctuations, which were captured during the CSF-TT. Even the recent survey on the practice trends of Indian physicians to manage iNPH did not capture what opening pressure cutoffs they used to diagnose iNPH.^15^ Similarly, in the systematic review by our group,^16^ it was found that each study had different ranges of opening pressures, and Wikkelsö et al. 1986 had included patients with a maximum CSF opening pressure of 30 cm H20 (22 mm Hg).^17^ Though the shunt responders had a higher CSF opening pressure as compared to non responders, this did not reach statistical significance, probably because of low sample sizes, which were not powered to detect these differences.

Our findings did not demonstrate a significant association between routine CSF protein levels and CSF-TT responsiveness. However, shunt responders had significantly higher CSF protein levels compared to non-responders. We did not perform CSF biomarker analysis, such as tau proteins; however, previous studies suggest that, beyond total protein, specific biomarkers, including t-tau and p-tau, may have a potential role in iNPH.^18^ According to a CSF biomarker study, elevated CSF t-tau and p-tau are linked to shunt non-responsiveness in iNPH, likely reflecting underlying neurodegeneration and poorer outcomes. In contrast, lower levels predict better postoperative gait and cognitive improvement. A meta-analysis confirmed that p-tau and t-tau, but not Aβ1-42, differentiate between shunt responders and non-responders. 7 Biomarker panels combining t-tau, Aβ40, and MCP-1 show promise for improving diagnostic accuracy and distinguishing iNPH from cognitive and movement disorders.^19^ We found that a narrower callosal angle was significantly associated with higher proteins. This finding has to be confirmed in larger cohorts.

Periventricular white matter changes are another biomarker, which likely reflects the raised intraventricular pressure leading to trans-ependymal pressure gradient. This leads to increased water content in the white matter in contrast to the ischemic white matter changes as seen on diffusion microstructure imaging sequences, and there is evidence to suggest that these patients respond better to surgery than those without. Increased water retention could be differentiated from white matter microstructural changes, and these imaging techniques should be studied systematically to generate new imaging biomarkers.^20^

This study has several limitations. First, it was conducted with a relatively small sample size at a single center, which may introduce bias and limit the generalizability of our findings. Second, as this was a hypothesis-generating study not originally designed for these outcomes, the observed association between higher CSF opening pressure and CSF-TT responsiveness, although statistically significant, requires validation in larger and more diverse cohorts.

Third, we relied on a single time-point measurement of CSF opening pressure. Continuous or repeated measurements of CSF pressure and physiology could provide greater insight into fluctuations and help establish cutoffs with optimal sensitivity and specificity. Similarly, CSF-TT responsiveness was assessed at a single 24-hour time point. Multiple assessments over time, with improvement defined at any of these intervals, may increase sensitivity, particularly for patients who exhibit delayed responses that our study may have missed.

Finally, there is no current diagnostic category for patients with imaging features of INPH and elevated opening pressures. This group may represent a distinct subgroup with prognostic differences compared to patients with lower pressures. Longitudinal studies with extended follow-up are needed to better define the natural history and clinical trajectory of these patients.

## Conclusion

Our study suggests that higher CSF opening pressure, even within accepted diagnostic ranges, is associated with better tap test responsiveness and may serve as a simple predictor of shunt responsiveness in iNPH. Routine CSF biochemical parameters were not predictive, though higher protein levels correlated with a narrower callosal angle on MRI. These findings support the role of integrating opening pressure with imaging features in the preoperative evaluation of iNPH and prognostication. Larger, prospective multicenter studies are warranted to investigate longitudinal changes in opening pressures, incorporating advanced CSF biomarkers and standardized imaging scales, to refine prognostication and improve patient selection for shunting.

## Data Availability

All data produced in the present work are contained in the manuscript

## Financial Disclosures for the previous 12 months

The authors declare that there are no additional disclosures to report.

## Ethical Compliance Statement

The study was reviewed and approved by the Institute Ethics Committee at the All India Institute of Medical Sciences, New Delhi (No. IECPG-222/20.4.23, RT-14/07.06.23). The participants and their legal guardians provided informed consent to participate in this study. We confirm that we have read the Journal’s position on issues involved in ethical publication and affirm that this work is consistent with those guidelines.

---

**Supplementary Table 1:** Comparison of CSF findings between shunt responders and non-responders.

| Parameter | Shunt responder (n=15) | Non-responder(n=9) | P value |
| --- | --- | --- | --- |
| Opening pressure median (IQR)<br>n=15 | 18(15-21) | 10(6-20) | 0.2 |
| Cell count/mm<sup>3</sup> median (IQR)<br>n=24 | 0(0-5) | 0(0-0) | 0.6 |
| CSF Protein mg/dL median (IQR)<br>n=24 | 56(50-78) | 25(10-34) | 0.001 |
| CSF Glucose mg/dL, mean (SD)<br>n=24 | 67.7(20.9) | 71.8(13.8) | 0.6 |
| Ratio of CSF: Blood glucose<br>mean(SD) n=18 | 0.6(0.1) | 0.6(0.2) | 0.4 |

**Supplementary Figure 1.**
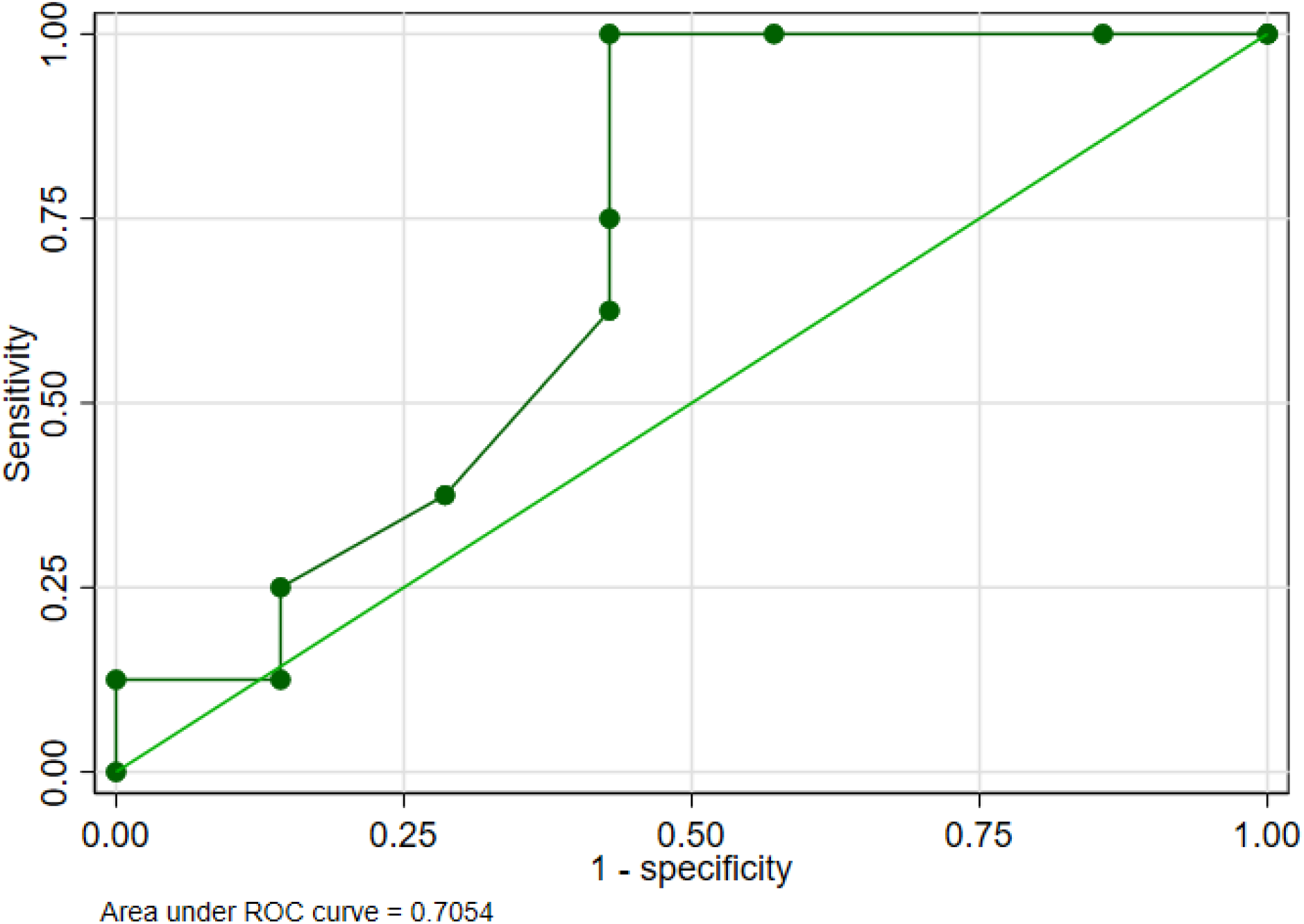
Receiver operating characteristic (ROC) curve for CSF opening pressure in predicting shunt responsiveness

